# Maternal deaths associated factors in the Conflict-Affected North West Region of Cameroon. Lessons from a cross-sectional survey

**DOI:** 10.64898/2026.06.10.26355370

**Authors:** Eneigheo Emmanuel Achuondou, Umeikai Wycliff Ayaba, Ayama Carine Kuma, Kinyuy Emmanuella Talla

## Abstract

**Background:** Maternal mortality is a significant ongoing global public health crisis, particularly in sub-Saharan Africa and conflict-affected regions. Cameroon’s maternal mortality ratio is high at 406 deaths per 100,000 live births, while the ongoing Anglophone conflict has further exacerbated maternal healthcare delivery in the North West Region (NWR). Despite the evidence-based interventions like partographs, obstetric kits, birth preparedness plans, and active management of the third stage of labour, implementation gaps persist across health facilities.

**Objective:** The study aimed to assess factors related to preventable maternal deaths in the NWR of Cameroon by exploring maternal health service usage, implementation of obstetric measures, demand-side challenges, accessibility barriers, and health system weaknesses.

**Methodology:** The study employed a quantitative descriptive cross-sectional survey design. Data was collected with structured questionnaires from postpartum women and healthcare workers in selected health facilities and catchment communities in the NWR. Also, a multistage sampling technique was adopted, and Cochran’s formula generated a sample size of 109 respondents. In addition, data were analysed with SPSS v. 27 and STATA v. 18 via descriptive and inferential statistics.

**Results:** In this study,while 70.64% of females attended at least 4 ANC visits, only 38.53% met WHO ANC adequacy requirements. Facility delivery was 96.33%, yet only 38.46% received completed delivery plans. Conflict-related challenges affected access, with 44.95% reporting insecurity-associated movement difficulties, while 44.95% reported increased transportation expenses due to the conflict. Near-miss complications were reported among 27.52% of participants. Delivery record reviews indicated that obstetric kits were utilised in 81.76% of deliveries, partographs were accessible in 86.49% of records but correctly filled in just 60.81%, while oxytocin administration was 95.95%. Integrated Health Centres showed poorer adherence with intrapartum interventions compared with District/Regional Hospitals (p < 0.05).

**Conclusion:** In the NWR, maternal mortality was associated with accessibility, interconnected demand-side, conflict-related, and health-system determinants. While utilization of some maternal interventions was high, major implementation gaps such as weak referral systems, insufficient BEmONC readiness, poor partograph compliance, and conflict disruptions, continually compromise neonatal and maternal outcomes. Strengthening lower-level facilities, enhancing emergency referral systems, and improving implementation of evidence-based obstetric interventions are crucial for minimising maternal mortality in the NWR.

## Introduction

Maternal mortality remains a global public health crisis in the 21st century. Despite decades of global commitments and health system investments, approximately 260,000 women died during or following pregnancy and after childbirth in 2023. ^1^ Likewise, most neonatal deaths in sub-Saharan Africa (SSA) are linked to intrapartum complications or complications of preterm delivery, many of which can be managed with timely access to functioning Emergency Obstetric and Newborn Care (EmONC) services.^2,3^ The global Maternal Mortality Ratio (MMR) was 197 deaths per 100,000 live births in 2023, a figure that, while representing progress from past decades, remains inconsistent with the SDG 3, target 1 of reducing global MMR to below 70 per 100,000 live births by 2030.^4^ Achieving this target demands an annual reduction rate of nearly 15%, a pace rarely reached at national level in low- and middle-income countries.^1^ Approximately 92% of all maternal deaths in 2023 occurred in low- and lower-middle-income countries. Sub-Saharan Africa (SSA) alone accounted for an estimated 70% (182,000 deaths) of the global total, the majority of which were preventable.1 The lifetime risk of maternal death in high-income countries is 1 in 7,933, compared to a devastating 1 in 66 in low-income countries, a 120-fold disparity and evidence of persistent global health inequity.^1^

An often-overlooked driver of maternal mortality is political conflict and institutional fragility. Conflict and fragility-related factors accounted for 61% of global maternal deaths in 2023, with the MMR in conflict-affected areas reaching 504 per 100,000 live births, more than 5 times the MMR of 99 per 100,000 in stable environments.^1^ Conflict disrupts the foundations of maternal health: it destroys healthcare infrastructure, displaces skilled health workers, disrupts medical supply chains, affects transport systems, and prevents women from reaching care, as seen in the conflict-affected regions of Central and West Africa.^5^ Most maternal deaths result from preventable set of direct obstetric complications. Severe postpartum haemorrhage (PPH), hypertensive disorders of pregnancy (pre-eclampsia and eclampsia), puerperal infection, obstructed labour, and complications of unsafe abortion put together accounted for about 75% of all maternal deaths.^1^ The medical interventions to prevent or manage each of these are well-established as follows: Oxytocin, administered within 1 minute of birth, effectively prevents PPH, the single largest killer of women in childbirth; Magnesium sulphate prevents eclamptic convulsions and significantly reduces mortality from pre-eclampsia; Clean delivery practices and antibiotics prevent and treat sepsis; and Skilled birth attendance with Emergency Obstetric and Newborn Care (EmONC) addresses obstructed labour and birth asphyxia.

The persistent gap between the availability of these evidence-based interventions and their actual implementation at the facility level constitutes an important health system failure that demands urgent investigation to ascertain these gaps. The event of Maternal death is often compounded by neonatal death. Most neonatal deaths in SSA are linked to intrapartum complications such as birth asphyxia, preterm birth complications, and neonatal infections, many of which are manageable with timely access to functioning EmONC services.^2,3^ Maternal and newborn health are inseparably linked: the same skilled birth attendant, the same functional facility, and the same obstetric kit that saves the mother is the first line of defense for the newborn. Consequently, every failure to implement effective obstetric care is simultaneously a failure of neonatal care. Cameroon occupies a concerning position in global maternal health statistics. The national MMR was estimated at 406 per 100,000 live births, which places Cameroon among the highest-burden countries in the world.^6^ Harsono et al (2024) identified maternal age, level of education, parity, place of delivery, health care provider qualifications, delays in reaching health facilities, inadequate prenatal care, unemployment, and distance to health facilities as critical determinants of mortality in Cameroon, all of which are preventable or manageable with appropriate care, tools, and trained providers.^7^ To improve maternal health, barriers that limit access to quality maternal health services must be identified and addressed at both the health system and societal levels. According to WHO, factors that prevent women from receiving or seeking care during pregnancy and childbirth are: (a) Health system failures such as poor quality of care, including disrespect, mistreatment and abuse, insufficient numbers of and inadequately trained health-care providers, including shortages of essential medical supplies. (b) Social determinants, including income, access to education, race and ethnicity. (c) Harmful gender norms and inequalities that result in a low prioritisation of the rights of women and girls, including their right to safe, quality, and affordable sexual and reproductive health services. (d) External factors contributing to instability and health system fragility, such as climate and humanitarian crises.^8^

The Three Delays Model highlights delays in seeking care, reaching care, and receiving adequate care as major contributors to maternal mortality.^9^ Within health facilities, poor monitoring of labour using the partograph, inadequate infection prevention practices, and improper storage of life-saving commodities, such as oxytocin, increase risk.^10,11^ These gaps can be addressed by providing standardised obstetric delivery kits to promote infection prevention, promoting correct and consistent use of the partograph, supporting birth preparedness via structured delivery plans, strengthening oxytocin storage and cold-chain management, and contributing funds for staff motivations. Cameroon has taken steps to address this burden, including the introduction and scale-up of PBF, scale-up training and deployment of skilled birth attendants (midwives and Gynecologists), creation of additional primary health facilities and referral hospitals, scale-up of obstetric kits, and the introduction of the health voucher. However, as demonstrated by studies in Adamaoua Region, Maroua health district, and Nkambe health District.^12^ The translation of policy intent into facility-level practice has been impeded by knowledge gaps, supply chain weaknesses, motivational deficits, human resource limitations, and inadequate supervision.^12^

The North West Region (NWR) of Cameroon presents a challenging health landscape. Since late 2016, the Anglophone crisis, an armed conflict between Ambazonian separatist groups and the Cameroonian government, has devastated the healthcare system of the NWR and the South West Region resulting in displacement of thousands of people including children and pregnant women, non-functional health facilities due to security threats, disruptions in transport systems and supply chains, pregnant women reportedly delivering in bushes or traveling for weeks through insecure roads to access care,^12^ critical shortages of skilled maternity care providers, who have either fled, been displaced, or are unable to reach facilities safely, and electricity outages lasting months to years in many communities undermining cold-chain management for essential medicines, including oxytocin and documented instances of women dying en route to the hospital or immediately after reaching facilities because of delayed referrals.^12^ Despite these ongoing challenges, the NWR has maintained a downward trend in maternal death since 2020, recording 46, 37, and 27 deaths respectively in 2023, 2024, and 2025 with 90.9% of the contributory factors related to the first delay.^13^ In 2026, 11 deaths have been recorded between 1 to 8 weeks,^13^ figures likely under representing community maternal deaths and deaths from other private and informal settings. A study of determinants of maternal death in Mezam Division, the NWR’s most populous division identified postpartum haemorrhage, unsafe abortion, and hypertensive disorders as the leading direct causes of maternal mortality in Mezam Division. ^14^

Against this backdrop, the NWR continues to implement obstetric kits and health voucher in health facilities, which are part of a national strategy to curb maternal and neonatal mortality. An implementation gap may indirectly affect the fight against preventable maternal and neonatal deaths. Similarly, the partogram, a simple and inexpensive graphical tool for monitoring labour progress endorsed by WHO, remains irregularly used; birth plans are not properly explained to pregnant women during antenatal care, oxytocin is frequently stored outside the required cold chain (2–8°C), and health worker’s motivation compromised by insecurity, irregular pay, and inadequate supervision.^15^ The culmination of these deficits in a conflict-affected region creates an imperative for evidence-based operational research. While several studies have examined aspects of maternal mortality and obstetric care at the subnational level in Cameroon, including in Adamaoua, Maroua, and Mezam Divisions, important operational knowledge gaps remain. First, no study has comprehensively examined the demand-side, supply-side, and broader health system factors contributing to the increasing trend in maternal deaths. Second, the impact of the Anglophone crisis on maternal mortality in the region has not been empirically quantified, particularly with regard to its implications for the availability and implementation of obstetric kits. Therefore, this study aims to determine the factors associated with increasing maternal deaths in the North West Region of Cameroon. Specifically, it seeks: i. To assess maternal service utilization and the demand-side, supply-side, and the broader health system factors associated with maternal deaths in the NWR. ii. To assess the utilization of OKPs and other recommended interventions (partograms, birth plans, life-saving medicines like oxytocin) for childbirth and its relationship to MMR in the NWR. iii. To identify the factors limiting and enabling the use of OKPs and other recommended interventions (partograms, birth plans, life-saving medicines like oxytocin) for childbirth and its relationship to MMR in the NWR. iv. To identify context-specific and operational strategies for the reduction of maternal and neonatal mortality in the NWR.

## Methods

### Research Design

This study adopted a quantitative descriptive cross-sectional survey design. The design was appropriate because the study sought to collect measurable data at one point in time on factors associated with increasing maternal deaths The quantitative design allowed the researcher to assess maternal health service utilisation, demand-side barriers, accessibility constraints, utilisation of OKP interventions, birth preparedness, oxytocin use, and conflict-related barriers using structured questionnaires. The usage of structured questionnaires enabled the collection of standardised responses from respondents and allowed statistical analysis of patterns, frequencies, percentages, and associations.

### Research Area and Population

The study was conducted in selected health facilities and their catchment communities in the North West Region of Cameroon. The region was selected because it has experienced persistent maternal health challenges within the context of the ongoing Anglophone crisis, which has affected health facility functionality, transport systems, supply chains, staff availability, referral pathways, and women’s access to maternal healthcare services. Within these facilities, participants included: Post-partum women who have delivered within the last 6 months and accessed services at the selected facilities; Health care providers involved in maternal health service delivery, including nurses, midwives, and doctors; and Facility managers involved in maternal health planning, supply management, and obstetric kit implementation.

### Inclusion Requirement

**Table 1:** Inclusion and Exclusion Criteria.

| Variable | Inclusion | Exclusion |
| --- | --- | --- |
| Health facilities | Public, private or confessional health Facilities providing delivery services | Health facilities in highly insecured localities where access is completely impossible even from field |
|  |  | staff. |
|  |  | Facilities without delivery services. |
| Health Workers | All cadre of staff conducting deliveries in selected health facilities and who provide informed consent | Staff not involved in maternity care |
|  |  | Staff on temporary rotation less than 1 month |
| Women | Women aged 15-49 years<br>Delivered within the last 6 months or Bereaved (Maternal death in the family/community)<br>Resident in the NWR for > 6 months<br>Provide assent/consent | Women too ill to participate in the study<br>Women who decline consent |

### Sample Size Determination

The sample size was determined using Cochran’s formula for estimating a single population proportion.^16^ The formula is:

n= *Z^2^pq/d*^2^

Where:

n = minimum sample size

Z = standard normal value at 95% confidence level = 1.96

p = estimated proportion = 0.50

q = 1 − p = 0.50

d = margin of error = 0.10

Substituting:

n=

n= (1.96)^2^ (0.50) (0.50)/(0.10)^2^

n= 3.8416×0.25/0.01

n= 0.9604/0.01

n= 96.04

Therefore, the minimum calculated sample size was about 96 respondents.

*n _adj_* = 96/1−0.12

*n _adj_* = 96/0.88 =109.09

Therefore, the final sample size was rounded to 109 respondents.

This sample size was considered adequate because the study was a quantitative cross-sectional survey conducted in a conflict-affected setting where access to respondents and health facilities was constrained by insecurity, mobility restrictions, and logistical challenges.

### Sources of Quantitative Data

**Table 2:**
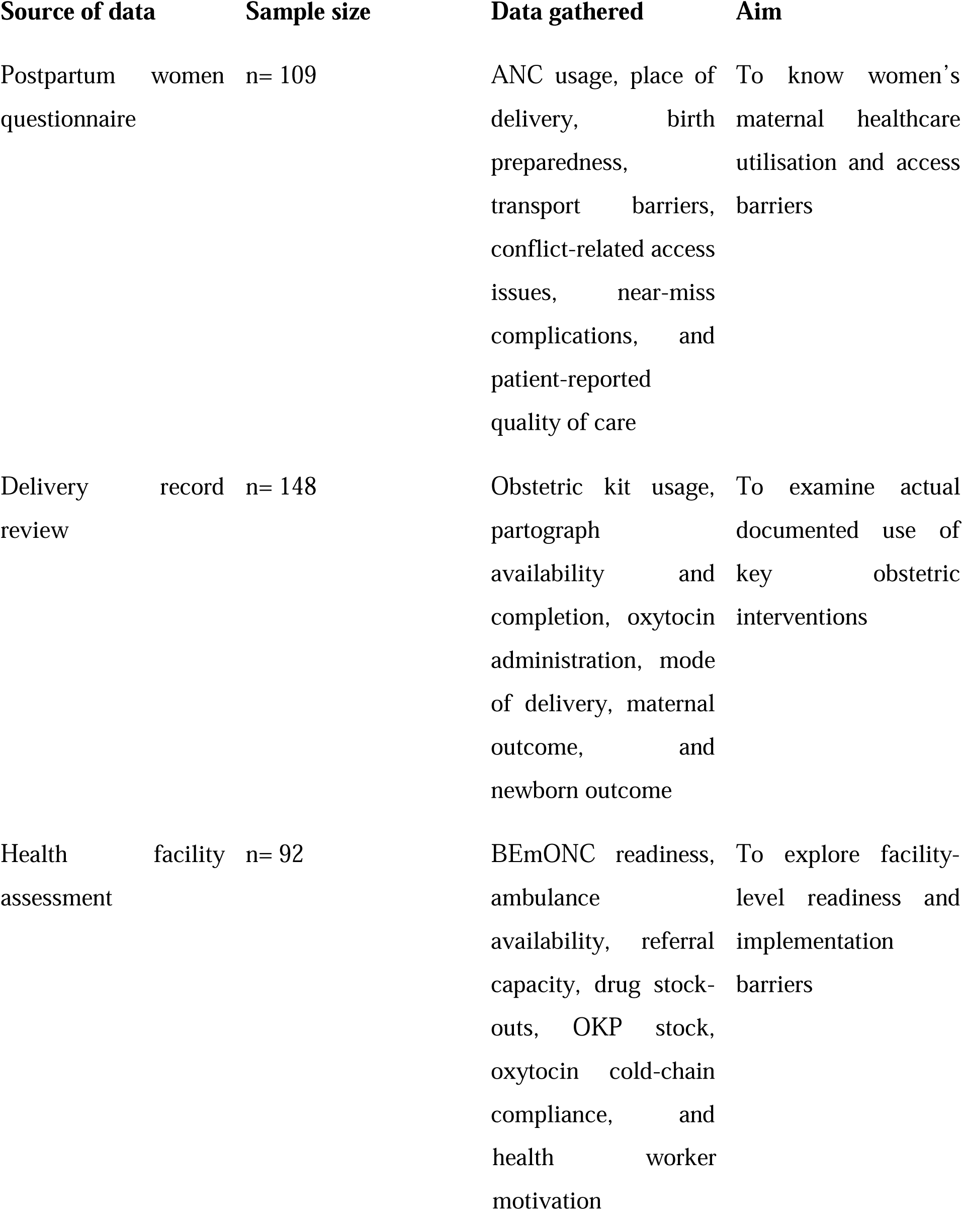
Description of Quantitative Data.

### Sampling Procedure

A multi-stage sampling approach was employed. Given the geographic, administrative, and security complexities of the NWR, a multistage sampling approach was appropriate for this study. The region is characterised by dispersed populations, varying levels of health service availability, and differing degrees of insecurity due to the ongoing crisis, which makes it difficult to implement a simple random or single-stage sampling strategy. Multistage sampling enabled the structural selection of study units at different levels; such as districts, health facilities, and individual participants; thereby improving feasibility while maintaining representativeness. At the first stage, districts were selected based on performance indicators (e.g., maternal mortality burden, service functionality, or security level), followed by the selection of health facilities within those districts. In subsequent stages, eligible participants, including post-partum women, bereaved families, near missess and health care providers, will be sampled within the selected facilities.

### Data Collection

Data for this study were collected using structured closed-ended questionnaires administered to postpartum women and healthcare workers in selected health facilities and catchment communities in the North West Region of Cameroon. The questionnaires were designed to capture quantitative information related to maternal health service utilization, antenatal care attendance, place of delivery, and delivery decision-making. Before the main survey, the questionnaire was pre-tested among 30 respondents in a non-selected health district to assess clarity, consistency, cultural appropriateness, and flow of questions. Necessary corrections and modifications were subsequently made. Trained research assistants administered the questionnaires over a two-week period to minimise incomplete responses and improve data accuracy. The training covered study objectives, ethical conduct, confidentiality, and questionnaire administration procedures. To ensure quality control, completed questionnaires were checked daily for completeness, consistency, and accuracy by the principal investigator before data entry and analysis.

### Method of Data Analysis

Data collected from the structured questionnaires were checked for completeness, coded, cleaned, and entered into SPSS V. 27 and STATA V. 18 for analysis. Descriptive and inferential statistical methods were employed in line with the study objectives and the nature of the variables collected. Descriptive statistics were first conducted to summarize socio-demographic characteristics, maternal health service utilization patterns, conflict-related barriers, facility characteristics, OKP implementation indicators, and quality-of-care variables. Frequencies, percentages, means, standard deviations, medians, interquartile ranges, and ranges were computed depending on the distribution and measurement level of variables. Composite indices were generated for selected multidimensional constructs, including the Conflict Exposure Index, BEmONC Readiness Score, and Oxytocin Cold Chain Compliance Score. These indices were subsequently categorized into levels such as low, moderate, high, partial, full, good, and poor based on predefined operational criteria. Bivariate analysis was performed to examine associations between categorical variables using Chi-square tests and Fisher’s exact tests where cell counts were small. Associations between intrapartum interventions and maternal outcomes, facility type and intervention compliance, and health worker satisfaction with OKP implementation practices were specifically examined. Statistical significance was set at p < 0.05. For continuous variables, measures of central tendency and dispersion were calculated. The analysed data were presented using tables, charts, frequencies, percentages, and narrative interpretation consistent with the study objectives.

## Result

### Socio-demographic Profile of Study Participants

**Table 3:**
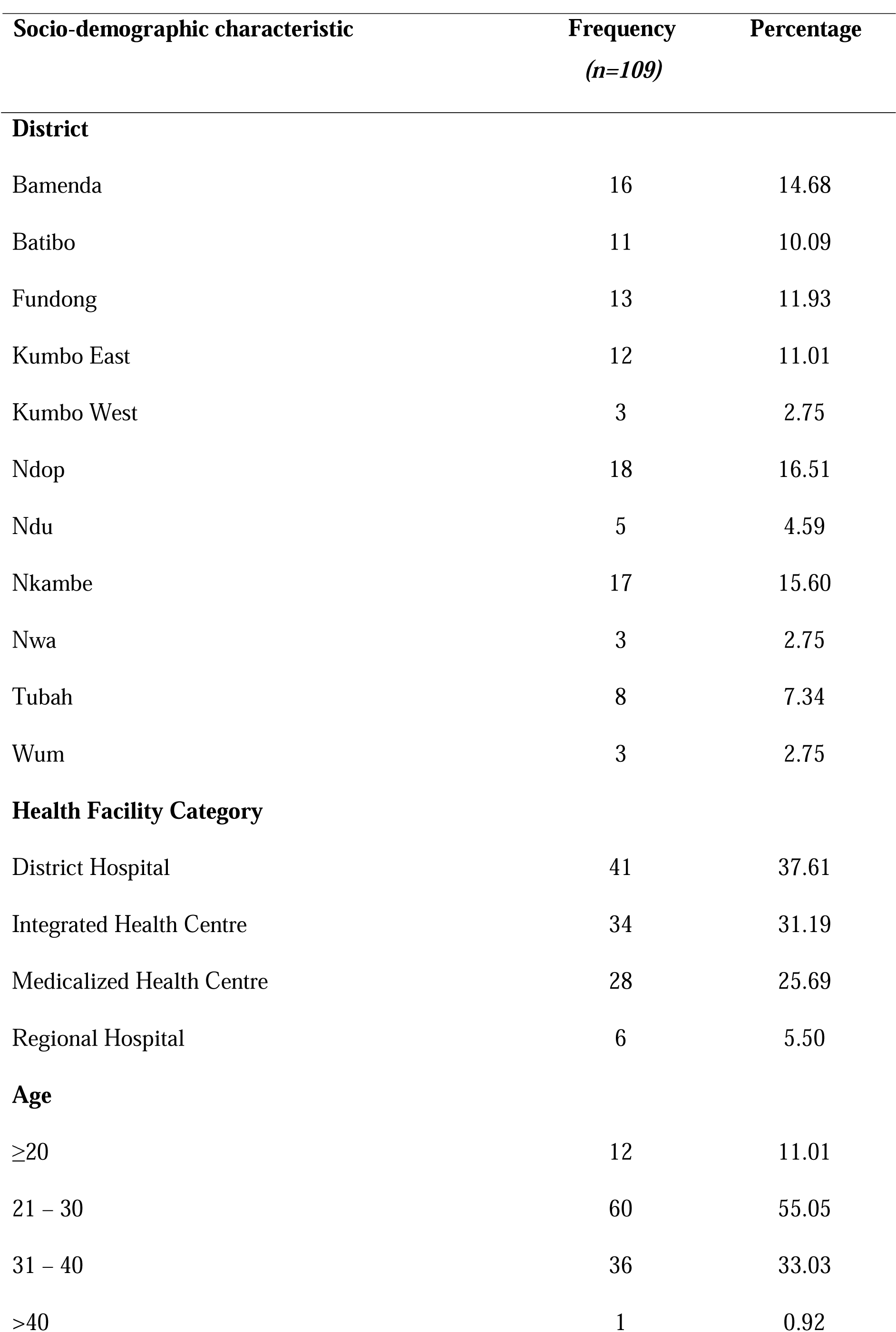
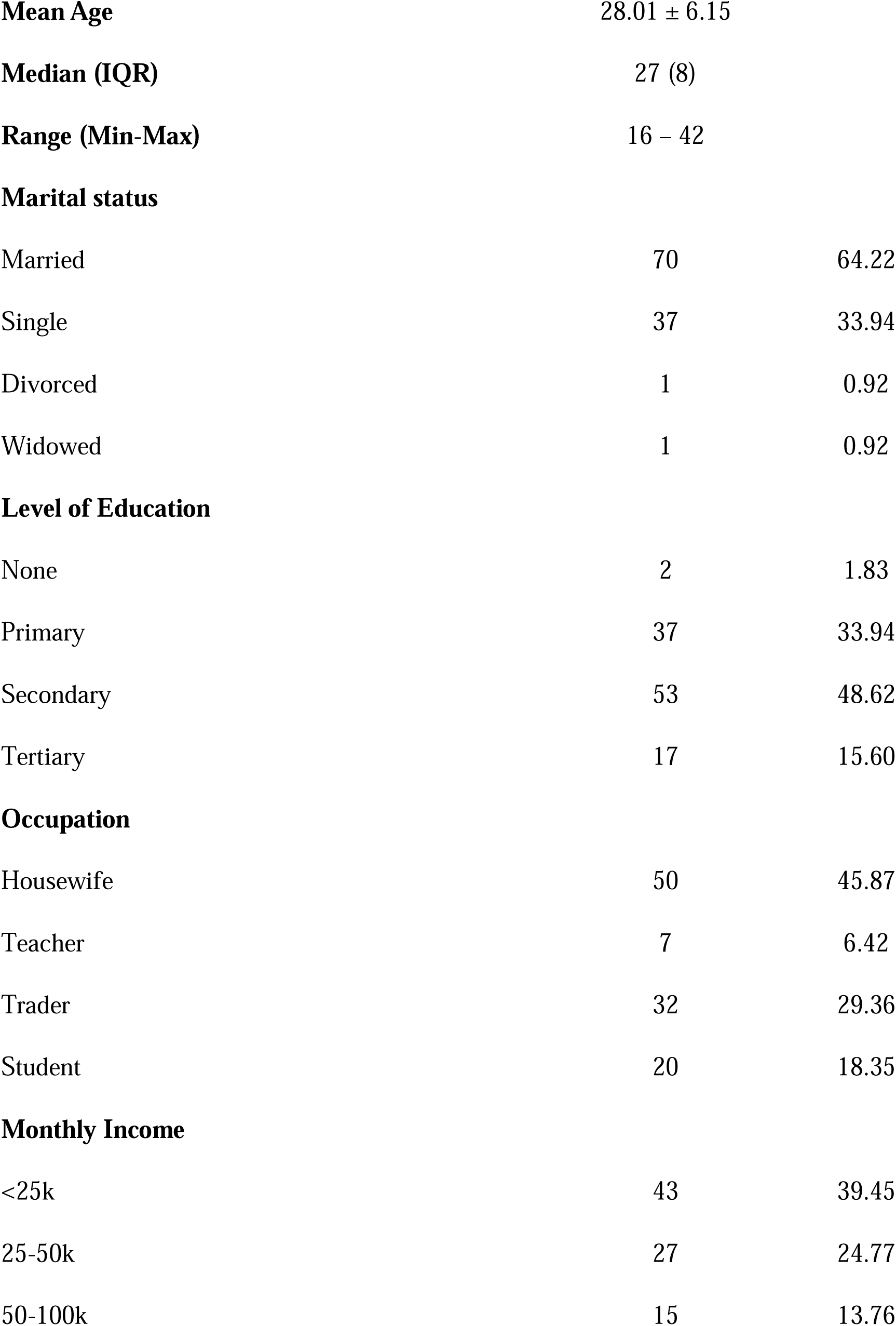
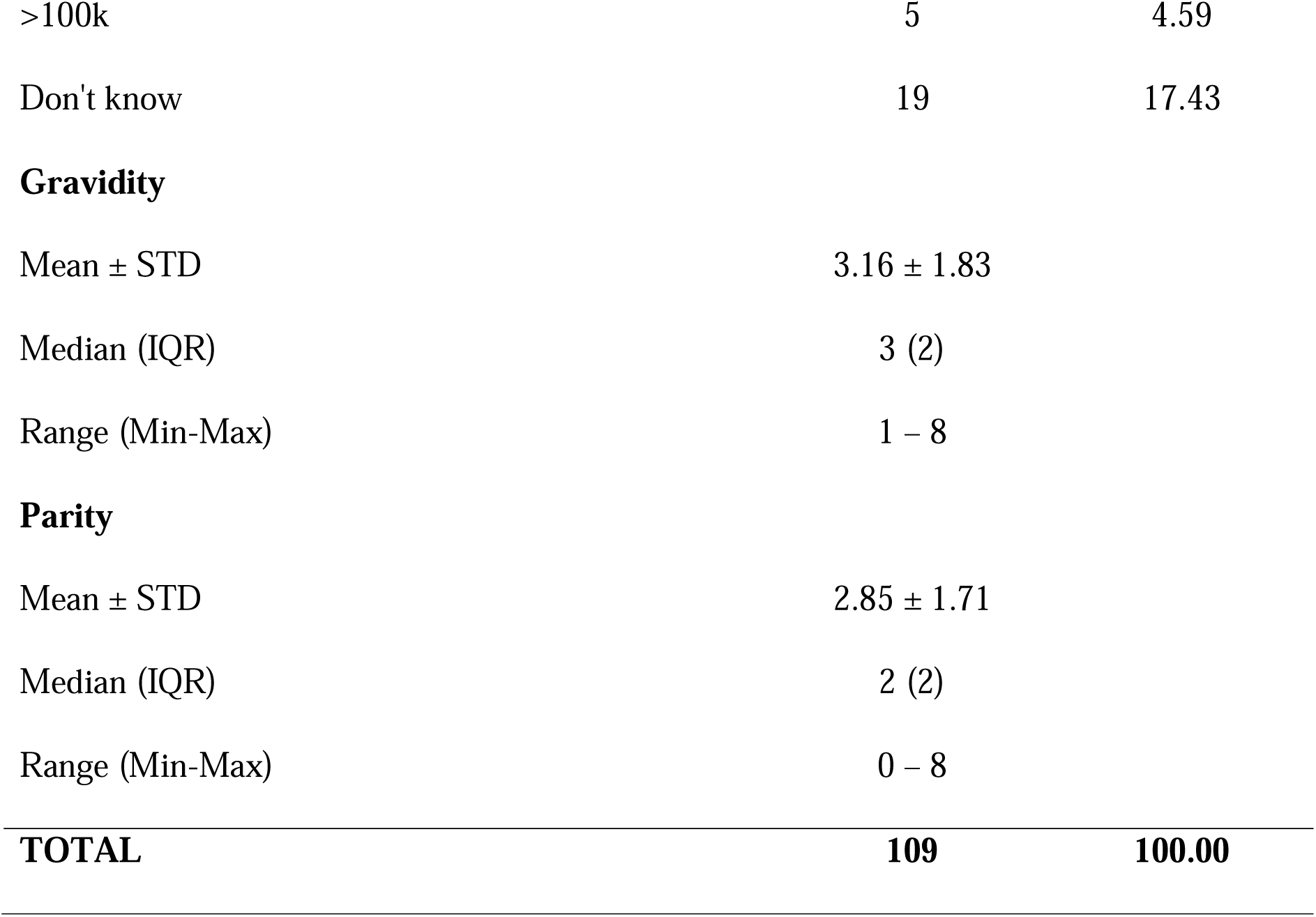
Socio-demographic characteristics of participants (n=109)

| **Socio-demographic characteristic** | **Frequency**<br>**(n=109)** | **Percentage** |
| --- | --- | --- |

| **District** |  |  |
| --- | --- | --- |
| Bamenda | 16 | 14.68 |
| Batibo | 11 | 10.09 |
| Fundong | 13 | 11.93 |
| Kumbo East | 12 | 11.01 |
| Kumbo West | 3 | 2.75 |
| Ndop | 18 | 16.51 |
| Ndu | 5 | 4.59 |
| Nkambe | 17 | 15.60 |
| Nwa | 3 | 2.75 |
| Tubah | 8 | 7.34 |
| Wum | 3 | 2.75 |
| **Health Facility Category** |  |  |
| District Hospital | 41 | 37.61 |
| Integrated Health Centre | 34 | 31.19 |
| Medicalized Health Centre | 28 | 25.69 |
| Regional Hospital | 6 | 5.50 |
| **Age** |  |  |
| ≥20 | 12 | 11.01 |
| 21 – 30 | 60 | 55.05 |
| 31 – 40 | 36 | 33.03 |
| >40 | 1 | 0.92 |
| **Mean Age** | 28.01 ± 6.15 |  |
| **Median (IQR)** | 27 (8) |  |
| **Range (Min-Max)** | 16 – 42 |  |
| **Marital status** |  |  |
| Married | 70 | 64.22 |
| Single | 37 | 33.94 |
| Divorced | 1 | 0.92 |
| Widowed | 1 | 0.92 |
| **Level of Education** |  |  |
| None | 2 | 1.83 |
| Primary | 37 | 33.94 |
| Secondary | 53 | 48.62 |
| Tertiary | 17 | 15.60 |
| **Occupation** |  |  |
| Housewife | 50 | 45.87 |
| Teacher | 7 | 6.42 |
| Trader | 32 | 29.36 |
| Student | 20 | 18.35 |
| **Monthly Income** |  |  |
| <25k | 43 | 39.45 |
| 25-50k | 27 | 24.77 |
| 50-100k | 15 | 13.76 |
| >100k | 5 | 4.59 |
| Don't know | 19 | 17.43 |
| **Gravidity** |  |  |
| Mean ± STD | 3.16 ± 1.83 | |
| Median (IQR) | 3 (2) |  |
| Range (Min-Max) | 1 – 8 |  |
| **Parity** |  |  |
| Mean ± STD | 2.85 ± 1.71 | |
| Median (IQR) | 2 (2) |  |
| Range (Min-Max) | 0 – 8 |  |
| **TOTAL** | **109** | **100.00** |

### Maternal Health Service Utilisation

**Table 4:**
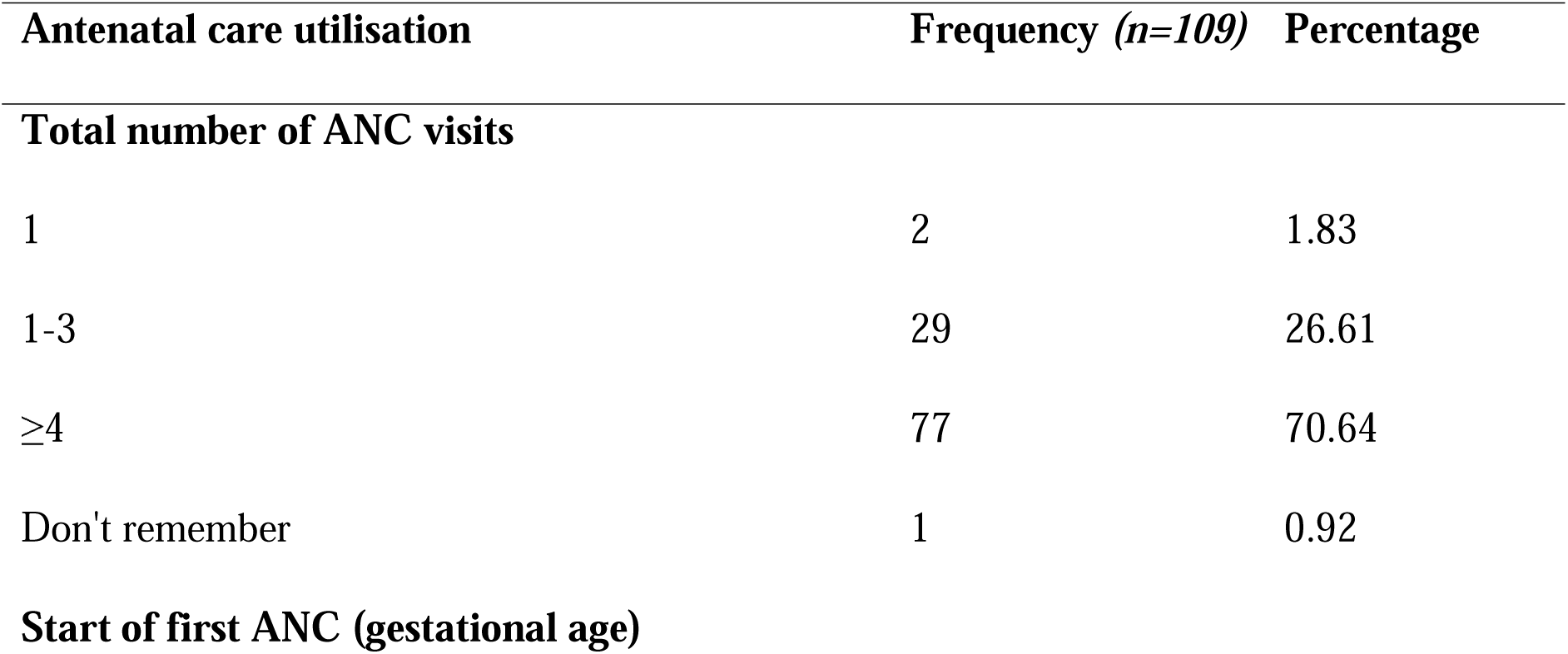
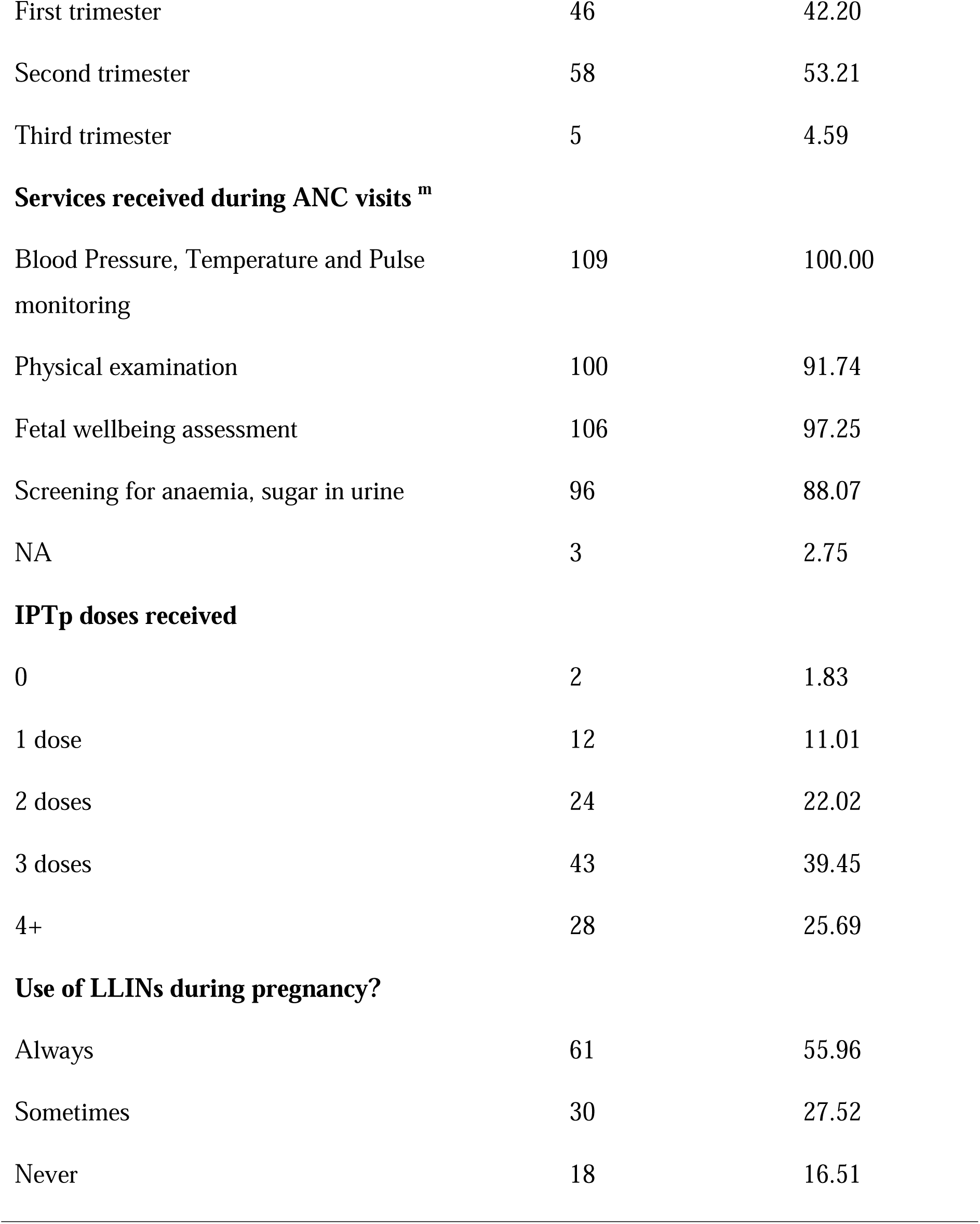
Antenatal care utilisation among participants (n=109)

**Table 5:** Proportion of participants meeting WHO ANC adequacy criteria (n=109)

| **Adequate ANC?** | **Frequency (n=109)** | **Percentage** |
| --- | --- | --- |
| No | 67 | 61.47 |
| Yes | 42 | 38.53 |

*Note: Adequate ANC is ANC visits greater than 4 and first ANC visit in 1<sup>st</sup> trimester*

**Table 6:** Place of delivery and birth attendant characteristics (n=109)

| **Variable** | **Frequency (n=109)** | **Percentage** |
| --- | --- | --- |
| **Place of delivery** |  |  |
| Facility | 105 | 96.33 |
| Home | 2 | 1.83 |
| TBA | 1 | 0.92 |
| On the way to the health facility | 1 | 0.92 |
| **If facility, type (n=105)** |  |  |
| Integrated Health Centre | 36 | 34.29 |
| Medicalized health center | 21 | 20.00 |
| District Hospital | 44 | 41.90 |
| Regional Hospital | 4 | 3.81 |
| **Who attended the delivery?** |  |  |
| Midwife | 44 | 40.37 |
| Nurse | 47 | 43.12 |
| Medical Doctor | 16 | 14.68 |
| Relative | 2 | 1.83 |

**Table 7:** Delivery plan receipt and birth preparedness counselling content (n=109)

| **Variable** | **Frequency (n=109)** | **Percentage** |
| --- | --- | --- |
| **Received filled delivery plan card during ANC?** |  |  |
| No | 64 | 61.54 |
| Yes | 40 | 38.46 |
| **If yes, was it explained one-on-one? n=40** |  |  |
| No | 4 | 10.00 |
| Yes | 36 | 90.00 |
| **Topics covered in the delivery plan <sup>m</sup>** |  |  |
| Expected Date of Delivery | 87 | 79.82 |
| Place of delivery | 68 | 62.39 |
| Danger signs | 89 | 81.65 |
| Transport | 55 | 50.46 |
| Emergency contact | 65 | 59.63 |
| Other topics | 40 | 36.70 |
| **Average time spent on delivery plan counseling with nurse** |  |  |
| <5 mins | 5 | 12.50 |
| 5-10 mins | 26 | 65.00 |
| >10 mins | 9 | 22.50 |

<sup>m</sup> indicates multiple response. Percentage shown for these variables are percentage of cases

**Table 8:** Delivery decision-making and autonomy among participants (n=109)

| **Variable** | **Frequency**<br>**(n=109)** | **Percentage** |
| --- | --- | --- |
| **Who decided where you delivered?** |  |  |
| Self | 70 | 64.22 |
| Husband | 21 | 19.27 |
| Mother-in-law | 1 | 0.92 |
| Health worker | 8 | 7.34 |
| Mother | 9 | 8.26 |
| **Main reason for the choice of delivery decision?** |  |  |
| Distance | 13 | 12.38 |
| Poor quality of health | 3 | 2.86 |
| Polite and receptive staff | 5 | 4.76 |
| Poor reception from health staff | 2 | 1.90 |
| Advice from health personnel | 2 | 1.90 |
| Good quality care | 80 | 76.19 |
| **Did husband/guardian refuse facility delivery?** |  |  |
| No | 90 | 82.57 |
| Yes | 2 | 1.83 |
| NA | 17 | 15.60 |

### Demand-Side and Accessibility Factors

**Table 9:**
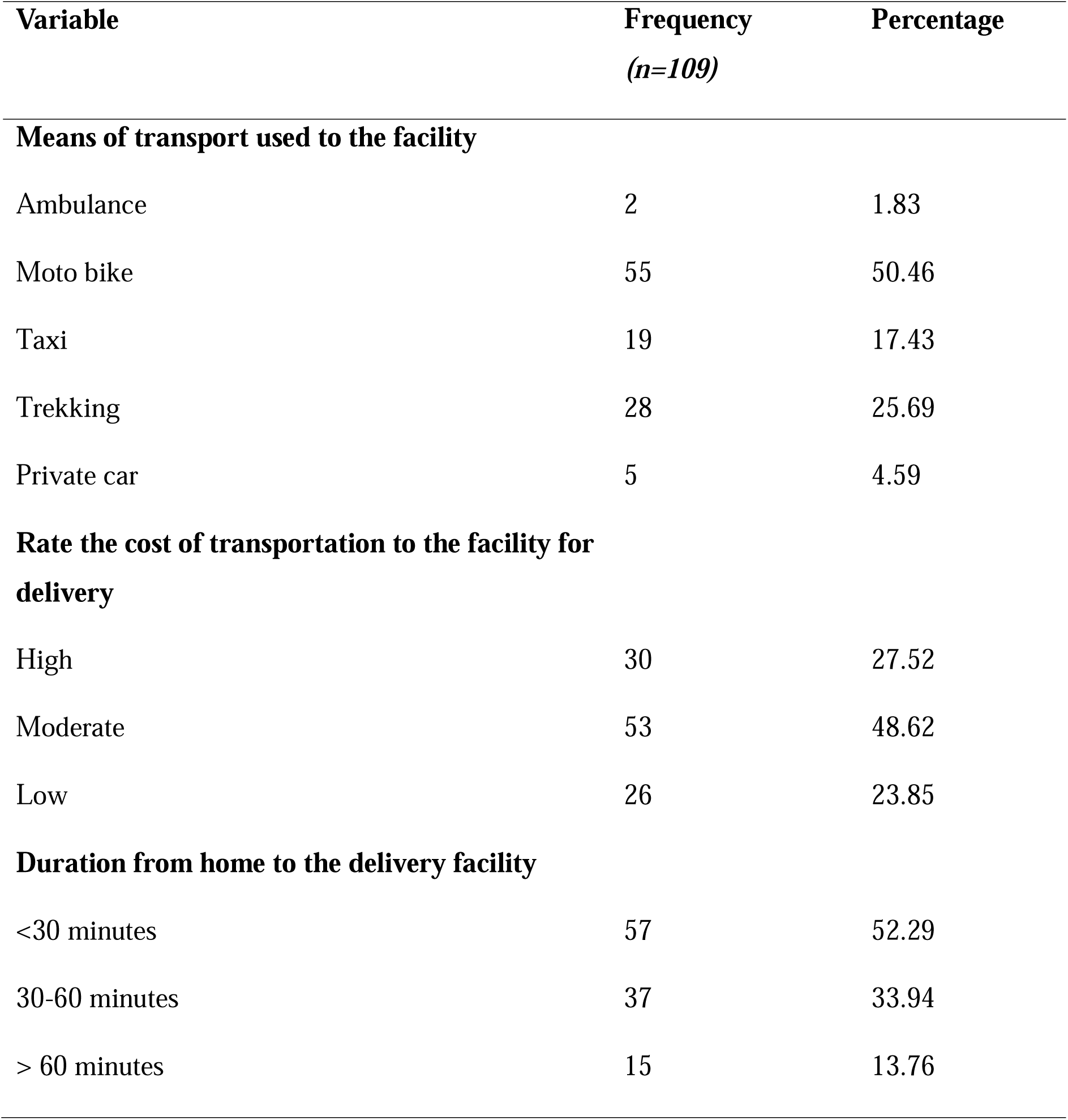
Transport characteristics and travel time to delivery facility (n=109)

| **Variable** | **Frequency**<br>**(n=109)** | **Percentage** |
| --- | --- | --- |
| **Means of transport used to the facility** |  |  |
| Ambulance | 2 | 1.83 |
| Moto bike | 55 | 50.46 |
| Taxi | 19 | 17.43 |
| Trekking | 28 | 25.69 |
| Private car | 5 | 4.59 |
| **Rate the cost of transportation to the facility for delivery** |  |  |
| High | 30 | 27.52 |
| Moderate | 53 | 48.62 |
| Low | 26 | 23.85 |
| **Duration from home to the delivery facility** |  |  |
| <30 minutes | 57 | 52.29 |
| 30-60 minutes | 37 | 33.94 |
| > 60 minutes | 15 | 13.76 |

**Table 10:** Conflict-related barriers to facility access during delivery (n=109)

| **Variable** | **Frequency**<br>**(n=109)** | **Percentage** |
| --- | --- | --- |
| **Insecurity/curfew make movement to the facility difficult at night** |  |  |
| Always | 28 | 25.69 |
| Sometimes | 49 | 44.95 |
| Never | 32 | 29.36 |
| **Fighting/violence prevented facility visits** |  |  |
| Always | 5 | 4.59 |
| Sometimes | 37 | 33.94 |
| Never | 67 | 61.47 |
| **Crisis increase the cost of transportation** |  |  |
| Always | 49 | 44.95 |
| Sometimes | 33 | 30.28 |
| Never | 27 | 24.77 |

**Table 11:** Near-miss complications experienced during pregnancy or delivery (n=109)

| **Experienced any of the following during pregnancy or delivery?** | **Frequency**<br>**(n=109)** | **Percentage** |
| --- | --- | --- |
| Heavy bleeding | 2 | 1.83 |
| Infection | 21 | 19.27 |
| Baby Resuscitation | 7 | 6.42 |

**Table 12:** Prevalence of near-miss complications (n=109)

| **Near-miss complications was** | **Frequency**<br>**(n=109)** | **Percentage** |
| --- | --- | --- |
| Present | 30 | 27.52 |
| Absent | 79 | 72.48 |

**Table 13:** Birth preparedness counselling received by women (n=109)

| **Birth preparedness counselling** | **Frequency**<br>**(n=109)** | **Percentage** |
| --- | --- | --- |
| **Where you counseled to move closer to the health facility towards expected date of delivery** |  |  |
| Yes | 64 | 58.72 |
| No | 45 | 41.28 |
| **Where you counseled to stay in the health facility for some time because of any danger sign in pregnancy** |  |  |
| Yes | 44 | 40.37 |
| No | 65 | 59.63 |

### Delivery Record Review

**Table 14:** Obstetric kit and partogram use in reviewed delivery records (n=148)

| **Variable** | **Frequency (n=148)** | **Percentage** |
| --- | --- | --- |
| **Health Facility Category** |  |  |
| District Hospital | 47 | 31.76 |
| Integrated Health Centre | 53 | 35.81 |
| Medicalized Health Centre | 37 | 25.00 |
| Regional Hospital | 11 | 7.43 |
| **Kit used?** |  |  |
| No | 27 | 18.24 |
| Yes | 121 | 81.76 |
| **Partogram present?** |  |  |
| No | 20 | 13.51 |
| Yes | 128 | 86.49 |
| **Partograph correctly filled?** |  |  |
| No | 58 | 39.19 |
| Yes | 90 | 60.81 |
| **Oxytocin administered?** |  |  |
| No | 6 | 4.05 |
| Yes | 142 | 95.95 |
| **Mode of delivery** |  |  |
| Caesarean section | 19 | 12.84 |
| Instrument delivery | 3 | 2.03 |
| Normal vaginal delivery | 126 | 85.14 |
| **TOTAL** | **148** | **100.00** |

**Table 15:** Maternal and neonatal outcomes from delivery record review (n=148)

| **Variable** | **Frequency (n=148)** | **Percentage** |
| --- | --- | --- |
| **Maternal Outcome** |  |  |
| Complications | 6 | 4.05 |
| Stable | 142 | 95.95 |
| **Child Outcome** |  |  |
| Alive | 146 | 98.65 |
| Stillbirth | 2 | 1.35 |

**Table 16:**
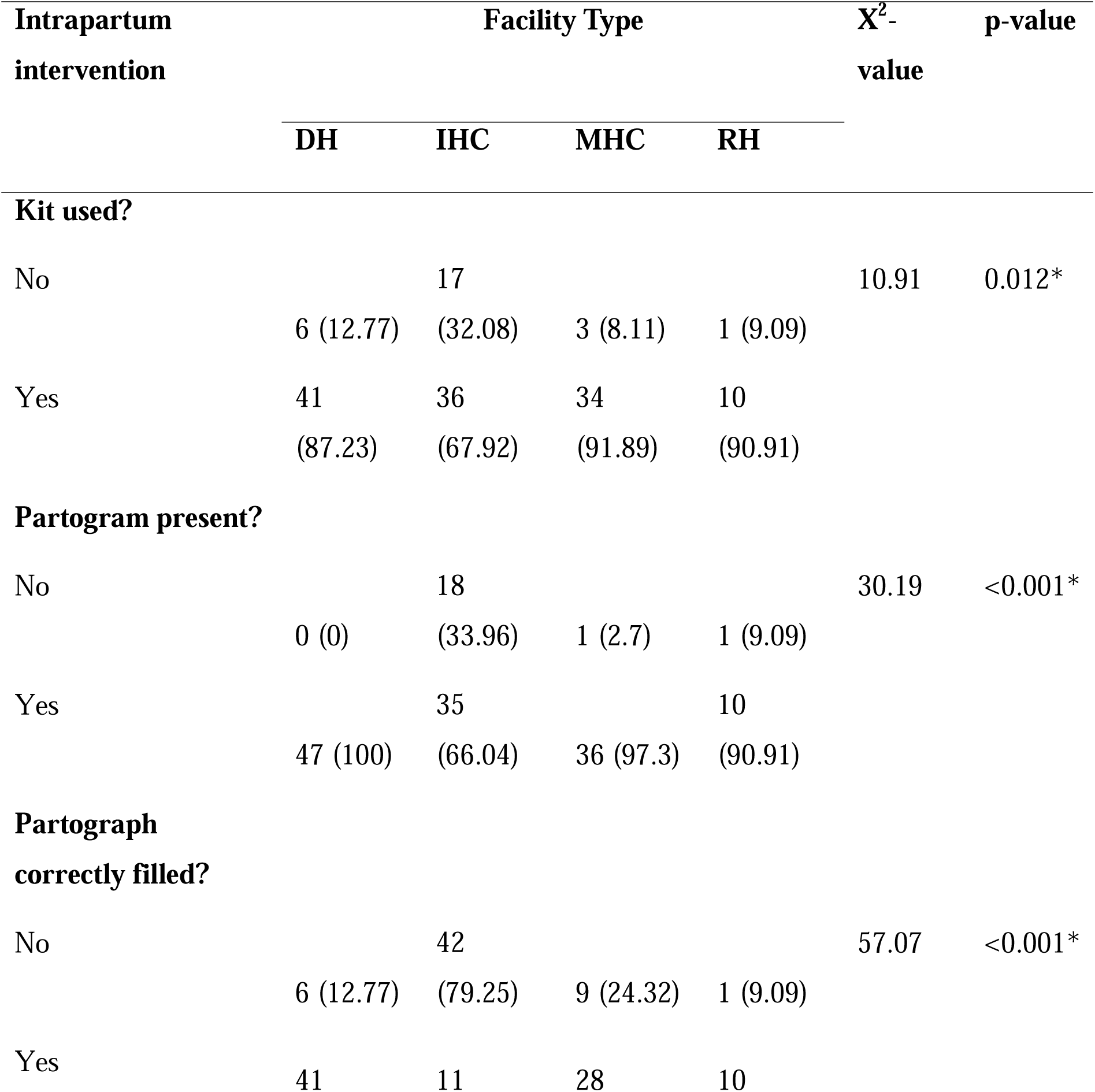
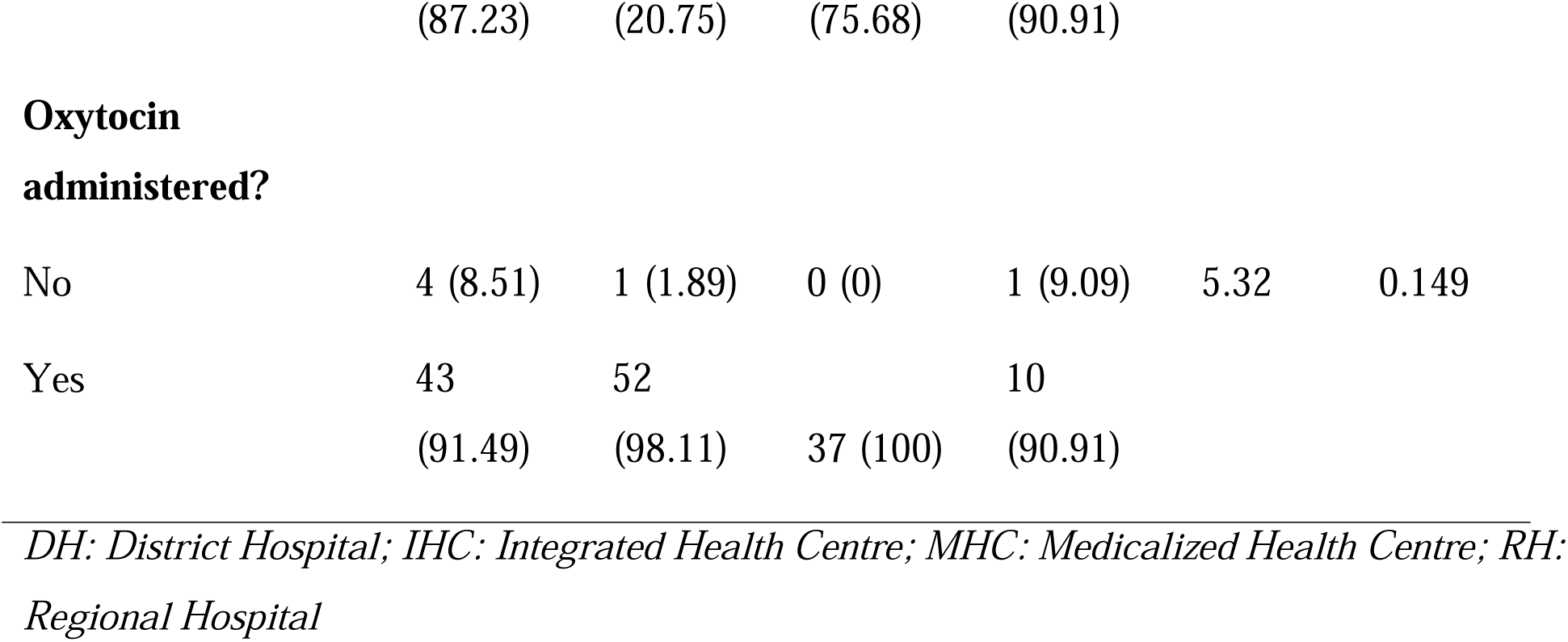
Intrapartum intervention compliance by facility type (n=148)

### Health Facility Infrastructure and Supply

**Table 17:** Facility-reported maternal and perinatal deaths and staffing levels (n=92)

| Variable | Mean | STD | Median | IQR | Range |
| --- | --- | --- | --- | --- | --- |
| Number of maternal deaths recorded in the past 1 year | 0.30 | 0.98 | 0 | 0 | 0 – 7 |
| Number of perinatal deaths recorded in the past 1 year | 3.55 | 7.12 | 1 | 3 | 0 – 31 |
| Number of midwives conducting delivery in the facility | 4.26 | 4.36 | 3 | 5 | 0 – 27 |
| Number of Medical doctors involve in Delivery and or C/S | 1.67 | 2.29 | 1 | 2 | 0 – 14 |

**Table 18:** Availability of BEmONC signal functions in health facilities (n=92)

| Variable | Frequency<br>(n=92) | Percentage |
| --- | --- | --- |
| **Oxytocin injection currently available?** |  |  |
| Yes | 89 | 100.00 |
| **MgSO4 injection currently available?** |  |  |
| No | 27 | 30.34 |
| Yes | 62 | 69.66 |
| **Manual vacuum aspiration kit available?** |  |  |
| No | 21 | 23.60 |
| Yes | 68 | 76.40 |
| **Newborn resuscitation kits available?** |  |  |
| No | 21 | 23.60 |
| Yes | 68 | 76.40 |
| **Blood transfusion service available?** |  |  |
| No | 29 | 32.58 |
| Yes | 60 | 67.42 |

**Table 19:** BEmONC readiness score and categorization (n=92)

| **BEmONC Readiness** | **Frequency (n=92)** | **Percentage** |
| --- | --- | --- |
| Mean ± STD | 3.77±1.56 |  |
| Median (IQR) | 4 (2) |  |
| Range (Min-Max) | 0 – 5 |  |
| Full | 45 | 48.91 |
| Inadequate | 20 | 21.74 |
| Partial | 27 | 29.35 |

*Note: BEmONC readiness score ranges from 0–5: oxytocin available + MgSO4 + MVA kit + newborn resus kit + blood transfusion. Categorization: Inadequate (0–2), Partial (3–4), Full (5)*

**Table 20:** Referral system capacity and functionality (n=92)

| Variable | Frequency (n=92) | Percentage |
| --- | --- | --- |
| **Functional Ambulance available?** |  |  |
| No | 67 | 75.28 |
| Yes | 22 | 24.72 |
| **Fuel for emergency transport available?** |  |  |
| Always | 17 | 19.10 |
| Never | 62 | 69.66 |
| Sometimes | 10 | 11.24 |
| **Referral success rate in the last 6 months** |  |  |
| 50-80% | 28 | 31.46 |
| <50% | 21 | 23.60 |
| >80% | 40 | 44.94 |

### OKP Utilisation Factors

**Table 21:**
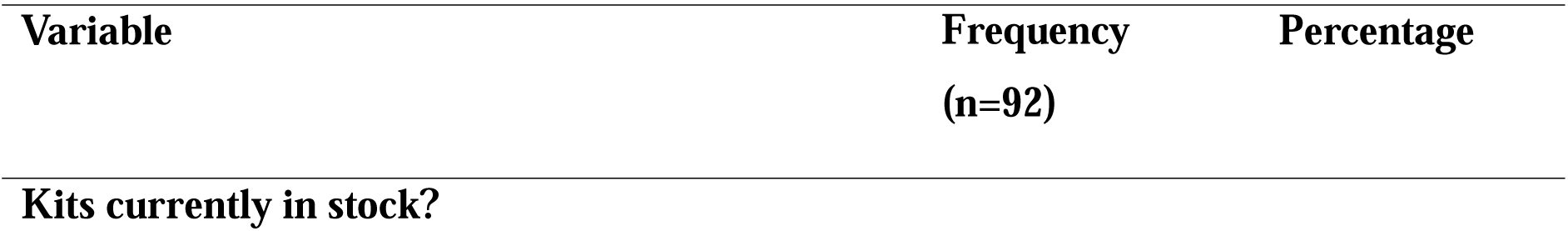
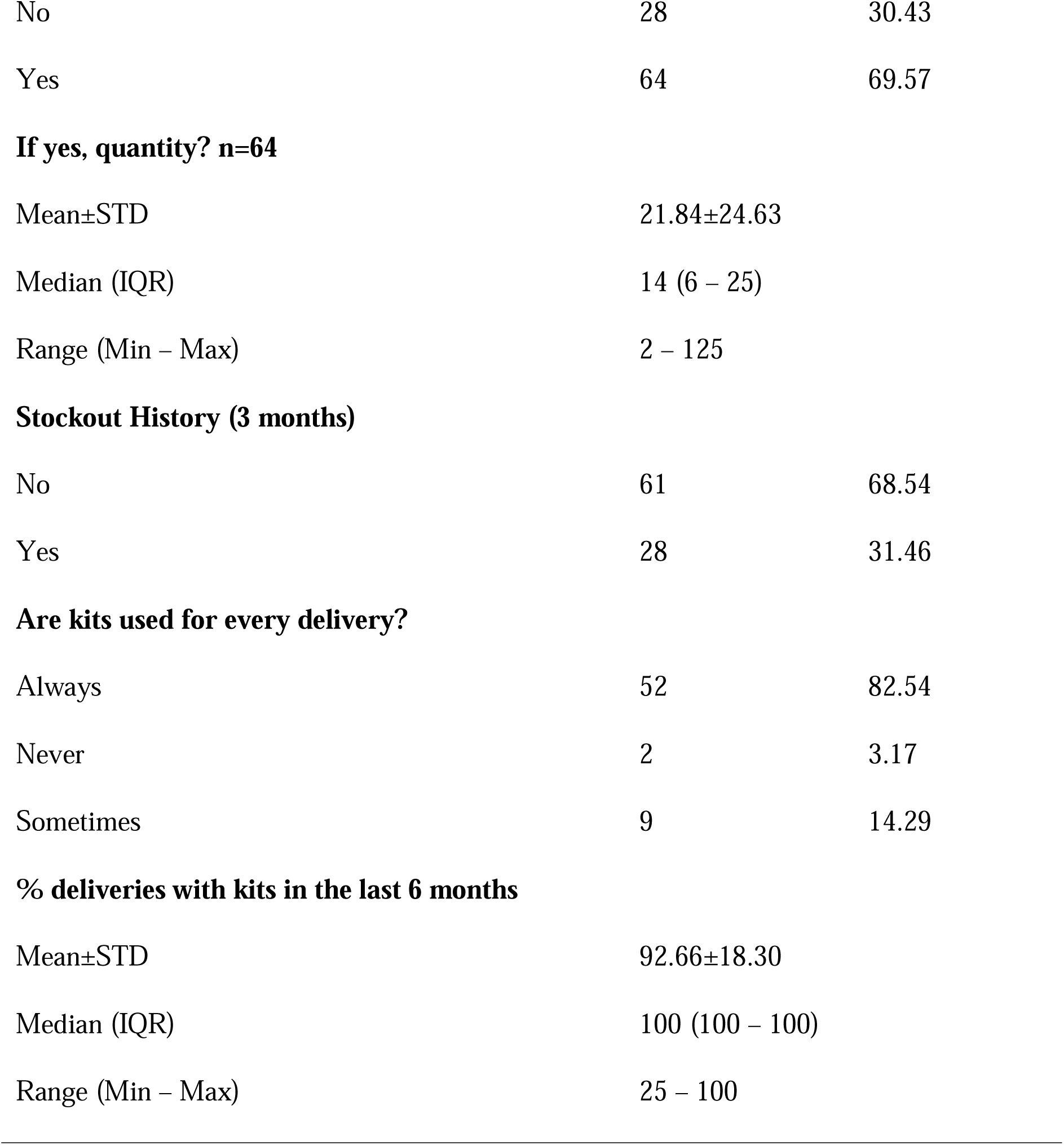
Obstetric kit stock availability and utilisation (n=92)

| Variable | Frequency<br>(n=92) | Percentage |
| --- | --- | --- |
| **Kits currently in stock?** |  |  |
| No | 28 | 30.43 |
| Yes | 64 | 69.57 |

**If yes, quantity? n=64**
|  |  |
| --- | --- |
| Mean±STD | 21.84±24.63 |
| Median (IQR) | 14 (6 – 25) |
| Range (Min – Max) | 2 – 125 |
| **Stockout History (3 months)** |  |  |
| No | 61 | 68.54 |
| Yes | 28 | 31.46 |
| **Are kits used for every delivery?** |  |  |
| Always | 52 | 82.54 |
| Never | 2 | 3.17 |
| Sometimes | 9 | 14.29 |

**% deliveries with kits in the last 6 months**
|  |  |
| --- | --- |
| Mean±STD | 92.66±18.30 |
| Median (IQR) | 100 (100 – 100) |
| Range (Min – Max) | 25 – 100 |

## Discussion

### Maternal Service Utilisation and Health System Determinants of Maternal Mortality

The results of this study demonstrate that maternal mortality in the North West Region is influenced by interconnected deficiencies in maternal solution utilisation, accessibility, emergency preparedness, and health system functionality within a conflict-impacted setting. While 70.64% of women attended at least four ANC visits, only 38.53% met the WHO criteria for sufficient ANC because most women initiated ANC during the second trimester. This finding suggests that the challenge in the North West Region is not merely ANC attendance, but delayed initiation of care, which limits early detection and management of pregnancy-related complications. This finding resonated with the research by Egbe et al (2016), who identified delayed utilisation of maternal health solutions and insufficient prenatal care as major determinants of maternal mortality in Mezam Division, Cameroon.^14^ Similarly, Gelaw et al (2024) explained that delays in seeking maternal healthcare substantially increase the risk of preventable maternal deaths in low-resource environments.^17^ In spite of poor ANC adequacy, facility delivery was remarkably big (96.33%), with most deliveries attended by nurses and midwives. Nevertheless, this big institutional delivery rate did not completely translate into sufficient maternal preparedness. Only 38.46% of women received a completed delivery plan during ANC, while 61.54% did not. This indicates weak execution of birth preparedness interventions in spite of relatively good facility utilisation. Insufficient delivery planning likely contributes to delays during obstetric emergencies, especially regarding transport arrangements, emergency decision-making, and referral preparedness. This finding is consistent with Feyisa Balcha et al (2024), which emphasised that birth preparedness plus complication readiness are important for reducing maternal mortality via early recognition of danger signs and timely care-seeking.^18^ Demand-side and accessibility barriers were shaped by the ongoing armed conflict. Over 70% of respondents reported that insecurity plus curfews impacted movement to health facilities either always or sometimes, while 75.23% stated that the crisis increased transportation costs. More so, 35.78% of women experienced big conflict exposure. These findings confirm that the Anglophone crisis has disrupted maternal healthcare access in the North West Region via restricted mobility, insecurity, and economic hardship. This finding resonated with the February 2026 report by the WHO, which stated that conflict-impacted environments account for the biggest burden of maternal deaths worldwide because insecurity disrupts transportation systems, access to facilities, and continuity of maternal healthcare solutions.^4^ Supply-side plus wider health system deficiencies were equally evident. Only 48.91% of facilities demonstrated full BEmONC readiness, while 75.28% lacked functional ambulances and 69.66% reported no consistent fuel availability for emergency transportation. These findings reveal major referral plus emergency response weaknesses likely contributing to preventable maternal deaths in the region. Also, insecurity accounted for 57.14% of staff absenteeism and desertion cases, further weakening maternal health solution delivery. This finding corresponds to the study by Zhang et al (2023), who reported that armed conflicts weaken maternal health systems via destruction of infrastructure, shortages of skilled personnel, disrupted supply chains, and minimised emergency referral capacity.^19^

### Utilisation of Obstetric Interventions and Maternal Mortality in the North West Region

The findings of this article demonstrate that utilisation of OKP components plus other recommended childbirth interventions in the North West Region was relatively big, While substantial execution gaps persisted regarding quality, consistency, and facility-level compliance. Delivery record review showed that obstetric kits were utilised in 81.76% of deliveries, partographs were present in 86.49% of records, and oxytocin was administered in 95.95% of deliveries. These findings imply improved execution of maternal health interventions compared to earlier regional monitoring reports, which documented only 32% OKP utilisation across facilities in the North West Region. Nevertheless, in spite of this relatively big utilisation, maternal complications were still reported in 4.05% of reviewed delivery records, which indicates that availability and usage alone do not necessarily translate into optimal maternal outcomes. A major concern identified in this article was the poor quality of partograph completion. While partographs were available in most records, only 60.81% were correctly filled. This finding is clinically important because incomplete labour monitoring restricts early detection of obstructed labour, fetal distress, plus prolonged labour, which are major contributors to postpartum haemorrhage, sepsis, and maternal mortality. More so, Integrated Health Centres demonstrated majorly poorer adherence with partograph availability and correct completion compared to District and Regional Hospitals (p<0.001). Similar findings were reported by Bedada et al (2020) who observed that poor adherence to partograph protocols remains common in low-resource environments in spite of WHO recommendations, largely due to workload pressures, insufficient supervision, plus restricted staff competency.^20^ The article also identified important deficiencies in birth preparedness execution. While 81.65% of women received counselling on danger signs during ANC, only 38.46% received a completed delivery plan card. This demonstrates weak operationalisation of structured birth preparedness interventions in spite of relatively sufficient counselling coverage. Birth preparedness planning is essential for minimising delays in seeking and reaching care during obstetric emergencies. According to Gebre et al (2015), individualised birth plans improve timely decision-making, emergency transportation arrangements, and referral preparedness, especially in fragile and conflict-impacted environments.^21^ While oxytocin utilisation was high, broader emergency obstetric readiness remained insufficient. Only 48.91% of facilities achieved full BEmONC readiness, while ⅓ lacked blood transfusion solutions plus magnesium sulphate availability. These deficiencies minimise the efficiency of OKPs and life-saving maternal interventions, thereby sustaining preventable maternal mortality risks within the North West Region.

### Factors Influencing the utilisation of Obstetric Interventions in the North West Region

The findings of this article revealed that utilisation of OKP components plus other recommended maternal health interventions in the North West Region was shaped by multiple interrelated health systems, infrastructural, workforce, and conflict-related factors. While obstetric kit utilisation (81.76%), partograph availability (86.49%), plus oxytocin administration (95.95%) were relatively big, execution quality and consistency remained suboptimal across facilities. The findings imply that availability of interventions alone is insufficient to guarantee effective maternal healthcare delivery in fragile and conflict-impacted environments. One of the major restricting factors identified was poor quality of execution, especially concerning partograph completion. While partographs were present in most delivery records, only 60.81% were correctly completed. More so, Integrated Health Centres recorded majorly poorer compliance with partograph usage and completion compared with District and Regional Hospitals (p<0.001). This finding implies important disparities in technical capacity and supervision across facility categories. Similar findings were reported by Zelellw and Tegegne (2018) who identified insufficient staff training, big workload, and poor supervision as major barriers to effective partograph utilisation in low-resource environments.^22^ Weak facility readiness also majorly restricted execution of recommended interventions. While oxytocin was available in every assessed facility, only 48.91% achieved full BEmONC readiness. About ⅓ of facilities lacked blood transfusion solutions and magnesium sulphate availability, while 23.60% lacked newborn resuscitation kits and manual vacuum aspiration kits. These deficiencies compromise the management of postpartum haemorrhage, eclampsia, septic abortion, plus neonatal asphyxia, which remain leading causes of maternal and neonatal mortality. According to Tembo et al (2017), availability of essential signal functions is fundamental for minimising preventable maternal deaths. Conflict-related disruptions equally emerged as major barriers to execution.^23^ Staff absenteeism or desertion was reported in 15.22% of facilities, with insecurity accounting for 57.14% of cases. The ongoing Anglophone crisis likely contributes to minimised staffing levels, increased workload, and disruption of supportive supervision systems. More so, transportation insecurity and curfews might impact commodity distribution plus referral systems, thereby indirectly limiting OKP execution. Nevertheless, several enabling factors were also identified. big oxytocin availability, widespread facility-based delivery, and relatively high obstetric kit utilisation suggest that maternal health interventions have become integrated into routine maternity care within several facilities. The major adherence observed in District and Regional Hospitals further indicates that facilities with stronger staffing, infrastructure, supervision, and operational capacity are more likely to effectively enforce recommended maternal interventions. These findings show that solidifying facility readiness, workforce stability, supervision, and conflict-resilient health systems is essential for enhancing utilisation of obstetric interventions and reducing maternal mortality in the region.

### Context-Specific Strategies for Reducing Maternal and Neonatal Mortality in the North West Region

The findings of this article underline the need for context-specific plus operationally feasible strategies tailored to the fragile and conflict-impacted realities of the North West Region. The results demonstrated that maternal and neonatal mortality in the environment is strongly associated with delays in accessing care, weak emergency obstetric readiness, inconsistent execution of recommended maternal interventions, and conflict-related disruptions impacting both communities and health facilities. As a result, minimising maternal and neonatal mortality in the North West Region needs integrated interventions targeting demand-side, supply-side, and broader health system constraints simultaneously. One major operational strategy emerging from the findings is strengthening birth preparedness and emergency referral systems. While 81.65% of women received counselling on danger signs during ANC, only 38.46% received a completed delivery plan card. Also, only 1.83% of women accessed ambulance transportation during labour, while 50.46% depended on motorbikes and 25.69% trekked to facilities. These findings indicate persistent first plus second delays in accessing care. Solidifying structured birth planning, community emergency transport systems, maternity waiting homes, and referral coordination mechanisms may majorly minimise delays during obstetric emergencies. Similar recommendations were proposed by Gelaw et al (2024) who emphasised that minimising delays in seeking and reaching care is fundamental for lowering maternal mortality in low-resource environments.^17^ The findings equally support the need to solidify BEmONC capacity across lower-level facilities. Only 48.91% of facilities achieved full BEmONC readiness, while Integrated Health Centres demonstrated majorly poorer readiness levels compared with District and Regional Hospitals (p<0.001). More so, about ⅓ of facilities lacked blood transfusion solutions plus magnesium sulphate availability. These deficiencies compromise management of postpartum haemorrhage, eclampsia, sepsis, and neonatal asphyxia. Expanding BEmONC signal functions, enhancing supply chain systems, and decentralizing emergency obstetric care to peripheral facilities are therefore vital operational priorities. Another good strategy involves enhancing quality of care via solidified supervision and workforce capacity building. While partographs were present in 86.49% of reviewed records, only 60.81% were correctly completed. Staff absenteeism connected to insecurity was equally reported in 15.22% of facilities. These findings imply the importance of regular in-service training, supportive supervision, staff motivation systems, and conflict-sensitive workforce retention measures. According to Olea-Ramirez et al (2024), competent health personnel, proper labour monitoring, plus uninterrupted availability of life-saving interventions are essential for reducing preventable maternal and neonatal deaths.^24^ Lastly, the current findings emphasise that maternal mortality reduction measures in the region must incorporate conflict-responsive health system strengthening. Big levels of insecurity, transportation difficulties, plus disruption of health solutions show that conventional maternal health interventions alone might be insufficient without broader investments in resilient healthcare delivery systems adapted to humanitarian environments.

## Conclusion

This article examined the factors associated with increasing maternal deaths in the North West Region of Cameroon in the context of a prolonged humanitarian crisis. The findings demonstrate that maternal and neonatal mortality in the region are caused by interconnected demand-side, supply-side, accessibility, plus wider health system weaknesses In spite of the availability of evidence-based maternal health interventions. While maternal health solution utilisation appeared relatively big, important execution and quality gaps persisted. Even though 70.64% of women attended at least 4 ANC visits, only 38.53% met WHO criteria for sufficient ANC because most initiated ANC late, mainly during the second trimester. Facility-based delivery was big at 96.33%, with nurses plus midwives conducting over 83% of deliveries. Nevertheless, major deficiencies remained in birth preparedness and emergency planning. Only 38.46% of women received a completed delivery plan card during ANC in spite of 81.65% receiving counselling on danger signs. The article further demonstrated that conflict-related barriers majorly impacted maternal healthcare access. More than 70% of women reported that insecurity plus the ongoing crisis increased transportation difficulties and costs, while only 1.83% accessed ambulance transport during labour. Motorbikes remained the commonest means of transportation (50.46%), and 25.69% of women trekked to health facilities during labour. These results reflect persistent first and second delays in accessing emergency obstetric care in the conflict-impacted environment. Utilisation of recommended maternal interventions was generally big but inconsistently implemented. Obstetric kits were utilised in 81.76% of reviewed deliveries, oxytocin administration reached 95.95%, and partographs were available in 86.49% of delivery records. Nevertheless, only 60.81% of partographs were correctly completed, while Integrated Health Centres demonstrated majorly poorer compliance with recommended interventions compared with District and Regional Hospitals. These results show that execution quality, instead of mere availability, remains a major challenge. Health system readiness for emergency obstetric and newborn care was also insufficient. While oxytocin was universally available, only 48.91% of facilities achieved full BEmONC readiness. About ⅓ of facilities lacked blood transfusion solutions and magnesium sulphate availability, while 75.28% had no functional ambulance. More so, 69.66% reported no consistent fuel availability for emergency referrals. These deficiencies majorly compromise management of postpartum haemorrhage, eclampsia, sepsis, and neonatal emergencies. The article also identified workforce and operational challenges limiting effective execution of obstetric interventions. Staff absenteeism was reported in 15.22% of facilities, mainly due to insecurity, while stock-outs, insufficient training, workload, plus demotivation impacted partograph and kit utilisation. Facilities with bigger staff satisfaction demonstrated majorly better OKP execution practices.

## Recommendations

a. Integrated Health Centres showed majorly poorer readiness plus intervention adherence compared with District and Regional Hospitals. Therefore, the regional Delegation of Public Health should prioritise upgrading lower-level facilities via provision of magnesium sulphate, blood transfusion solutions, newborn resuscitation kits, manual vacuum aspiration kits, plus emergency obstetric equipment to enhance management of postpartum haemorrhage, eclampsia, sepsis, and neonatal complications.

b. Only 24.72% of facilities had functional ambulances, while 69.66% reported no regular fuel availability for emergency referrals. Thus, government plus humanitarian partners need to establish conflict-adapted emergency referral networks, community ambulance systems, fuel support mechanisms, and maternity waiting homes for women from remote and insecure communities to minimise delays in accessing emergency obstetric care.

c. While partographs were available in 86.49% of delivery records, only 60.81% were correctly completed. Hence, continuous in-service training, mentorship, routine clinical audits, and supportive supervision need to be implemented across facilities to improve compliance with WHO intrapartum monitoring standards and correct utilisation of obstetric kits.

d. Only 38.46% of women received a completed delivery plan in spite of big ANC attendance. Therefore, health facilities should institutionalise structured one-on-one birth preparedness counselling during ANC, while concentrating on transport arrangements, emergency contacts, recognition of danger signs, plus referral preparedness, especially for women living in conflict-impacted and hard-to-reach areas.

e. Health worker dissatisfaction, insecurity-related absenteeism, and perceived irregular payment of kit incentives negatively impacted execution of maternal interventions. As a result, the Ministry of Public Health should solidify staff motivation via regular payment of incentives, psychosocial support, security-sensitive deployment strategies, continuous professional development, plus enhanced supervision in conflict-impacted facilities.

Consequently, future research or studies should explore the efficiency of these interventions, particularly conflict-adapted referral models, staff incentive systems, plus BEmONC strengthening in minimising maternal and neonatal mortality.

## Study Limitations

i. The article employed a cross-sectional design, which restricted the ability to establish causal associations between identified factors plus maternal mortality outcomes.
ii. Due to the ongoing Anglophone crisis, some insecure health areas and inaccessible societies could not be included in the current research. This might have restricted representation of the most severely impacted populations plus facilities.
iii. Some information collected from postpartum females, including ANC attendance, delivery experiences, and complications, depended on self-reporting plus participant recall. This might have introduced recall bias.
iv. The article depended majorly on facility-based records and accessible participants. Maternal deaths happening in communities, informal environments, or non-functional facilities during the crisis might not have been fully captured. This results in possible underestimation of the true burden of maternal mortality within the North West areas.

## What is already known on this topic

What is already known on this topic

Maternal mortality is still high in conflict-impacted sub-Saharan Africa. Also, conflict disrupts access to maternal healthcare and care of emergency obstetric.

## What this study adds

The utilisation of Obstetric kit and adherence of partograph remain inconsistent in the NWR of Cameroon. Also, conflict-associated transportation challenges impact access to maternal healthcare. Lastly, lower-level facilities show weaker BEmONC readiness.

## Contributors

### Funding

None

## Ethics approval and Patient Consent

The full protocol was reviewed and approved by the Regional Delegation of Public Health and the North West Regional Fund for Health Promotion. An agreement in principle was obtained from the RDPH and from the respective districts involved in the study. Ethical clearance was obtained from the North West Regional Ethical Review Committee. Community authorizations was obtained from the traditional leaders in the study communities. Verbal informed consent was obtained from every participant before the survey. The voluntary nature of participation and the right to withdraw at any time was emphasized. All participant information was kept strictly confidential. No names or personal identifiers appeared in any database or report. Data was presented in aggregate form. The survey carried minimal risk. Health facilities in insecured locations was not sampled. Enumerators was selected from within the district of origin. While there was no direct benefit to participants, the study is aimed to benefit public health policy.

## Ethical Approval and consent to participate

Ethical clearance was obtained from the research ethics committee of the Faculty of Health Sciences, the University of Bamenda, with registration number 2026/0860H/UBa/IRB. In addition, verbal informed consent was obtained from each participant before data collection.

## Declaration

All authors have contributed to the research article, have reviewed and approved the final version, and consent to its publication. We confirm that this work is original and has not been submitted for publication elsewhere.

## Data availability declaration

The data generated for this research article is available on request from.

## Authors’ contributions

Eneigheo Emmanuel Achuondou initiated and led the writing of the paper. U.W.Ayaba, A.C. Kuma, and K.E. Talla reviewed the drafts and contributed to the intellectual content and writing of the paper. E.E. Achuondou, U.W.Ayaba, A.C. Kuma, and K.E. Talla contributed to the data analysis and discussion

